# Trajectories over time in the prevalence of substance and behavioral addictions and common mental health issues in Israel, 2022-2025

**DOI:** 10.64898/2026.02.16.26346376

**Authors:** Dvora Shmulewitz, Maor Daniel Levitin, Vera Skvirsky, Merav Vider, Shaul Lev-Ran, Mario Mikulincer

## Abstract

**Background:** Traumatic events, such as terror attacks and war, are expected to impact mental health. These potential effects can be explored by assessing the mental health of the general population of Israel, from before the events of October 7, 2023 and over the course of the Swords of Iron war.

**Methods:** General population data were collected from Jewish adults in Israel before October 7 (April 2022), after October 7 (December 2023), and over the course of the ongoing war (March 2024, June 2024, February 2025), in a series of repeated cross-sectional samples with longitudinal data on a subset of the respondents. Among a subset of the sample including individuals who were surveyed in April 2022 and at least one follow-up time point (N=1,368), we used regression analysis to model trajectories over time in prevalence of problematic non-medical use of alcohol, tobacco, cannabis, sedatives, prescription stimulants, and prescription opioid painkillers; problematic use of internet, social media, electronic gaming, gambling, pornography, and compulsive sexual behavior; and post-traumatic stress disorder (PTSD), depression, and anxiety. Trajectories were modeled overall and moderation analysis was used to determine if trajectories differed by gender or age.

**Results:** Different patterns were observed by outcome. Different trajectories were observed before and after December 2023, suggesting that the events of October 7^th^ and the early war may have been a key transition point, for problematic use of alcohol, tobacco, sedatives, opioid painkillers, gambling, sexual behaviors, internet and PTSD. Smooth changes over time were observed for problematic use of gaming and social media, and anxiety and depression. No changes over time were observed for problematic use of cannabis, stimulants, and pornography. Many outcomes showed different trajectories by gender and age.

**Conclusions:** Findings suggest possible effects of ongoing trauma and war and suggest that outcome-specific and group-specific strategies may be warranted. Monitoring the prevalence of addictions and other common mental health issues in the general population during and after nationally traumatic events is important to understand the evolving mental health of the population and provide information and resources for potential interventions. Awareness of the potentially harmful effects of such life events, as well as the consequences of maladaptive coping styles on health and well-being, should be increased.

## Introduction

Disasters, such as terror attacks and war, are known to increase risk for mental health disorders, such as post-traumatic stress disorder (PTSD), depression, and anxiety, as well as substance use and other addictive behaviors^1–6^. On October 7, 2023, Israel suffered one of the most severe mass casualty terror attacks in modern history, perpetrated by Hamas-led militants, with greater than 1200 fatalities, 9000 injured and 250 hostages taken^7,8^. The terror attacks led to an ongoing war, with life-threatening situations and continuing traumatic exposures, such as missile attacks from multiple fronts, and ongoing stressors. The breadth and scope of these events necessitates exploration of changes over time in the psychopathology of the general population in Israel, to add information about the effects of ongoing terror and war on mental health^9,10^.

As reviewed^11^, in the adult general population of Israel, these trauma exposures were associated with increased post-traumatic stress symptoms, depression, anxiety, substance use, and problematic gambling^12–20,21^. Over time, psychological distress may be decreasing somewhat^16,22,23^, but still remains prevalent. Additionally, women, those with more trauma exposure, and younger individuals were at higher risk for psychopathology^12–14,16–19,24–27^. Yet, most studies were limited in that they did not address longer term changes, from before the October 7^th^ attacks and over the course of the war. Furthermore, no studies have explored a wide range of problematic substance use or other addictive behaviors.

At the Israel Center for Addiction and Mental Health, we have data from an ongoing epidemiology study in the general population of adult Jews in Israel^28,29^, from April 2022, December 2023, March 2024, June 2024, and February 2025. These data can be used to explore changes in the prevalence of problematic substance use (tobacco, alcohol, cannabis, and prescription sedatives, stimulants, and opioid painkillers) and other potentially addictive behaviors (gambling, gaming, sexual behavior, pornography, social media, and internet use) and common mental health issues (PTSD, depression, anxiety) over time, from before October 7^th^ and during the war. For each outcome, the aims of this study were to: (1) explore if and how prevalence changed over time, overall; and (2) determine if the trajectory over time differed by gender or age. This information is important for understanding the possible mental health consequences of ongoing terror and war to identify areas of focus for efficacious intervention.

## Methods

### Sample and procedures

Data were collected from a general population sample of Jewish adults in Israel. Cross-sectional data were collected in April 2022^30^ (Apr 24-May 10), December 2023 (Nov 26-Dec 12)^28^, and February 2025 (Feb 5-Feb 24), with similar methodology, and repeated measures from a subset of the samples at subsequent timeframes (Supplemental Table 1). Respondents were recruited from a diverse panel of individuals who choose to participate in surveys (iPanel)^31^. Respondents were Hebrew speaking and Jewish, since substantial adaptations would be required to include different cultural groups^32^, and aged 18-70, as older individuals are less likely to participate in online surveys.

These cross-sectional samples were designed to be quasi-representative, i.e. to match the adult, Hebrew-speaking, Jewish population in Israel on prevalence of age, gender, geographic area, and religiosity. Therefore, sampling was done using specified quotas^33^, based on those socio-demographic variables. Quotas were set based on Israel Census Bureau data for the relevant years; deviations of up to 3% from the quotas were allowed. Potential participants were selected in two ways: all respondents surveyed previously were invited to participate, as were a random sample of other panel members. Individuals who accepted the invitation were screened against the quotas until the target numbers were met; if a specific quota was already filled, that individual was not asked to continue the survey. Two additional datasets were collected only from those who participated in December 2023: in March 2024 (Mar 6-31) and June 2024 (June 17-July 25)^29^.

All participants provided electronic informed consent. Confidentiality was maintained by iPanel not having access to survey responses, and identifying information not being available to the researchers. Survey methodology was consistent with the ICC/ESOMAR International Code on Market and Social Research^31^. The Institutional Review Board of the Reichman University approved the study.

The online survey, which assessed socio-demographics, substance use, addictive behaviors, psychopathology, and risk and protective factors, utilizing widely-used and valid measures, was conducted via Qualtrics^34^. Anonymous online surveys may be better for collecting potentially sensitive information such as substance use^35^. Participants received online gift cards worth ∼20 ILS for completing each survey. Quality assurance was maintained by: inviting registered individuals; 4-5 attention checks; and removing incomplete surveys. Supplemental Table 2 provides information on survey response and completion. Since this study explored changes over time, our analytical sample included 1,368 participants who were surveyed at Time 1 (T1: April 2022) and at least one additional timepoint (T2: December 2023; T3: March 2024; T4: June 2024; T5: February 2025).

### Measures

#### Problematic substance use

The Alcohol, Smoking and Substance Involvement Screening Test (ASSIST 3.1)^36,37^ was used. For each substance ever used non-medically (tobacco, alcohol, cannabis, sedatives, prescription stimulants, prescription opioid painkillers, and other drugs), respondents answered six questions related to the frequency of use, craving, and consequences of use. Responses were weighted and summed into substance involvement scores^36^. A binary variable for problematic use was defined as moderate (score of ≥11 or more for alcohol, ≥4 for other substances) or high (≥27) risk levels. These cut-offs were shown to have concurrent and discriminant validity, and are considered parallel to substance abuse (moderate) and dependence (high)^36,37^.

#### Problematic Gambling

The Problem Gambling Severity Index (PGSI)^38,39^ includes nine items assessing the frequency of gambling behaviors in the past 12 months, with response options from never (0) to almost always (3). The 9 items were summed and a binary variable indicating problematic gambling was created based on a sum of ≥5, the cut-off that led to the best discriminant validity and definition of gambling problems^39,40^.

#### Problematic Sexual behavior

The Bergen-Yale Sex Addiction Scale (BYSAS)^41^ includes 6 items assessing frequency of sexual behaviors, with response options from very rarely (0) to very often (4). A binary variable indicating problematic sexual behavior was positive if a response of “often” or higher was endorsed for more than half (≥4) of the items^41^.

#### Problematic Pornography Use

The Problematic Pornography Use Scale (PPUS)^42^ includes 12 items assessing statements about pornography use within the past year, with response options from never true (0) to almost always true (5). A binary variable indicating problematic pornography use was positive if a response of “often true” or higher was endorsed for more than half (≥7) of the items, similar to the definition of binary problematic sexual behavior.

#### Problematic Electronic Gaming

The Game Addiction Scale (GAS)^43,44^ includes 7 items assessing the frequency of gaming behaviors in the past 6 months, with response options from never (1) to very often (5). A binary variable indicating problematic gaming was positive if a response of “often” or higher was endorsed for more than half (≥4) of the items, as suggested^43^, as for the BYSAS and PPUS.

#### Problematic Social Media Use

The Bergen Social Media Addiction Scale (BSMAS)^45,46^ includes 6 items assessing frequency of social media behaviors in the past 12 months, with response options ranging from very rarely (1) to very often (5). A binary variable indicating problematic social media use was positive if a response of “often” or higher was endorsed for more than half (≥4) of the items, as for some other behavioral measures.

#### Problematic Internet Use

The Internet Addiction Test (IAT)^47,48^ includes 20 items assessing frequency of internet use behaviors in the past month, with response options ranging from not relevant (0) to always (5). All items were summed; a binary variable indicating problematic internet use was positive for sums ≥50, as in^49^.

#### Post-traumatic Stress Disorder (PTSD)

The Posttraumatic Stress Disorder Checklist – DSM-5 version (PCL-5)^50–52^ includes 20 items assessing how much respondent was bothered by PTSD-related problems in the past month, with response options from not at all (0) to extremely (4). A binary variable indicating potential PTSD was created based on the DSM-5 diagnostic rule, requiring rating of at least 1 B item, 1 C item, 2 D items, and 2 E items at a score of 2 (moderately) or above^50,52^. At T1, items were assessed from any traumatic event experienced. For T2-T5, items were assessed due to the attacks of October 7^th^ and the subsequent war. Note that since the war trauma was ongoing, these experiences may not be “post”-trauma, so we use the term PTSD more loosely, to indicate a high level of symptoms due to trauma.

#### General Anxiety

The Generalized Anxiety Disorder-7 (GAD-7)^53^ includes 7 items assessing how many days respondent was bothered by anxiety-related problems over the past two weeks, with response options from not at all (0) to nearly every day (3). All items were summed; a binary variable indicating potential anxiety was positive for sums ≥10. This measure was not assessed in April 2022.

#### Depression

The Patient Health Questionnaire-9 (PHQ-9)^54^ includes 9 items assessing how many days respondent was bothered by depression-related problems in the past two weeks, with response options from not at all (0) to nearly every day (3). All items were summed; a binary variable indicating potential depression was positive for sums ≥15. This measure was not assessed in April 2022.

#### Sociodemographics

Sociodemographic variables that were collected similarly over all timeframes included: gender, age, religiosity, area of residence, educational attainment, marital status, ethnicity, and socio-economic status (Supplemental Table 3). Gender and age were considered moderators of change over time.

### Statistical analysis

#### Missing data

Analyses were conducted on 1,368 participants who had data at T1 and at least one additional time point. This retention strategy ensured meaningful trajectory modeling while allowing for missingness across waves. Missing data analysis revealed 16.01% overall missingness across the longitudinal dataset, with 9 distinct missingness patterns identified through Little’s MCAR test^55^. We employed multiple imputation (MI) to handle missing data while preserving the uncertainty introduced by missingness, using the *mice* package in R^56^, with 16 imputed datasets, each with 10 iterations. Analysis was carried out within each imputed set, and results were averaged over all sets. The imputation process used predictive mean matching for continuous variables and logistic regression for categorical variables, with the predictor matrix excluding participant ID from both prediction and imputation. The validity of the imputation procedure was confirmed by successful preservation of binary outcomes distribution, comparing pre- and post-imputation data (Supplementary Figure 1).

#### Trajectory modeling framework

To examine changes in addiction and mental health outcomes from before the October 7, 2023 attacks through the ongoing conflict period, we employed a comprehensive trajectory modeling approach^57^. As T2 represents the immediate post-October 7 period marking the onset of war, and T2-T5 capture the prolonged conflict period, we tested six trajectory models for each outcome: (1) Intercept-only model: no change across time (null hypothesis); (2) Linear model: a constant rate of change across all five time points; (3) Quadratic model: acceleration or deceleration in change rates; (4) Cubic model: non-linear patterns with multiple inflection points; (5) Piecewise linear model: distinguishing between pre-war to immediate post-trauma change (T1-T2 as phase 1) and subsequent war period (T2-T5 as phase 2), with linear change in phase 2; and (6) Piecewise quadratic model: similar two-phase structure, with quadratic change in phase 2. For the piecewise models, phase 1 was coded as max(0, min(2, Time) - 1) and phase 2 as max(0, Time - 2), with phase2_sq representing the quadratic term for phase 2 (model 6). Models were estimated using mixed generalized linear models (GLM) with a binomial family and a logit link, using robust estimation, with the *lme4*, *sandwich*, and *clubSandwich* R packages^58–61^. Data on depression and anxiety were not available in T1, so trajectories were modeled from T2-T5, using models 1-4. For ease of presentation and interpretation, time points were modeled as equally spaced.

#### Model Selection Strategy

Model selection of the best fitting model (from the 6 listed above) employed a multi-faceted ensemble approach to overcome the conservative nature of single information criteria in small samples with limited time points. Five selection strategies were implemented: (1) Hierarchical F-tests: sequential testing from simpler to more complex models using likelihood ratio tests (α=0.05); (2) Adjusted R-squared prioritization: selecting models with highest adjusted R², requiring minimum 5% improvement over intercept; (3) Practical significance evaluation: combining effect size considerations (minimum 2% prevalence change) with statistical significance; (4) Leave-one-out cross-validation: assessing predictive accuracy through iterative model fitting; and (5) Ensemble voting: majority vote across all strategies with hierarchical selection as a tiebreaker. This comprehensive approach balanced model parsimony with adequate capture of meaningful change patterns.

#### Moderation Analyses

Moderation analysis was carried out to determine whether the trajectory of change differed by moderators (gender, age). For each outcome, we fitted each of the 6 trajectory models with moderator main effects and moderator × time interaction terms. For piecewise models, separate interactions were estimated for each phase to determine whether the moderator differentially influenced pre-war versus during-war trajectories. When multiple trajectory models showed interaction, the best-fitting model was selected, as done previously. For gender (categorical), to facilitate interpretation of effects, effect coding was used (Men = -0.5, Women = 0.5). Moderation significance was assessed through likelihood ratio tests comparing nested models with and without interaction terms, with False Discovery Rate (FDR^62^) correction at 10% to balance Type I and Type II error rates, given the exploratory nature of moderation analyses^63^. Age (continuous) was centered at the sample mean and analyzed using generalized linear models (GLMs) with binomial family rather than aggregated approaches, preserving maximal information and statistical power. Given the use of GLMs, likelihood ratio tests based on deviance differences were employed, with manual calculation as a fallback when standard ANOVA procedures encountered convergence issues. Significant moderation effects were probed by examining simple slopes or trajectories for both gender categories and ±1 SD from mean age. This approach provided interpretable estimates of how trajectories differed across demographic subgroups while maintaining appropriate statistical rigor.

## Results

### Sample descriptives

Of the sample (at T1), about half were women, secular, lived in Tel Aviv/Central area, had academic post-high school education; one-third were between ages 18-34; 38% were of Middle Eastern/North African ethnicity; and 62% were married (Supplemental Table 3). Prevalences of outcomes at all time points are shown in Supplemental Figure 1.

### Trajectories over time

The ensemble model selection approach identified distinct trajectory patterns across the outcomes (Table 1, Figure 1). Of these, 8 outcomes demonstrated two-phase trajectories with breakpoints after the October 7 attacks (problematic use of: tobacco, alcohol, sedatives, opioids, gambling, sexual behaviors, internet; and PTSD), while 4 exhibited continuous change (problematic use of: gaming, social media; anxiety and depression) and 3 showed stable patterns (problematic use of: cannabis, stimulants, pornography).

**Table 1.** Trajectory models and changes from pre-war (T1, 2022) to present (T5, 2025)

| Outcome | Best-Fitting Model | Pseudo R<sup>2</sup> | p-value<sup>a</sup> | T1 Prevalence (%) | T2 Prevalence (%) | T5 Prevalence (%) | T1→T2 Change (%) | Overall Change (%) | Trajectory Pattern |
| --- | --- | --- | --- | --- | --- | --- | --- | --- | --- |
| Two-Phase Trajectories (Breakpoint at Oct 7) |  |  |  |  |  |  |  |  |  |
| Problematic tobacco use | Piecewise linear | 0.996 | 0.004 | 37.6 | 33.6 | 33.0 | -3.9 | -4.6 | Immediate decrease, stabilized |
| Problematic alcohol use | Piecewise linear | 0.846 | 0.154 | 12.4 | 11.2 | 16.1 | -1.2 | +3.7 | Initial decrease, then increase |
| Problematic sedative use | Piecewise linear | 0.948 | 0.052 | 4.4 | 9.7 | 10.4 | +5.3 | +6.0 | Sharp increase, continued rise |
| Problematic opioid use | Piecewise quadratic | 0.999 | 0.030 | 2.9 | 4.6 | 5.3 | +1.7 | +2.3 | Increase, accelerating |
| Problematic gambling | Piecewise quadratic | 1.000 | <0.001 | 5.0 | 6.7 | 6.0 | +1.7 | +1.0 | Initial increase, curved |
| Problematic sexual behaviors | Piecewise linear | 0.979 | 0.021 | 9.6 | 3.7 | 3.4 | -5.9 | -6.3 | Sharp decrease, continued decline |
| Problematic internet use | Piecewise linear | 0.962 | 0.038 | 6.4 | 8.2 | 7.7 | +1.8 | +1.3 | Immediate increase, plateau during conflict |
| PTSD | Piecewise linear | 0.894 | 0.106 | 11.5 | 22.9 | 12.8 | +11.4 | +1.3 | Sharp spike immediately post-trauma, gradual decline |
| **Continuous Trajectory** |  |  |  |  |  |  |  |  |  |
| Problematic Gaming | Quadratic | 0.954 | 0.046 | 2.4 | 3.6 | 3.5 | +1.2 | +1.1 | Gradual curved increase |
| Problematic social media use | Cubic | 0.998 | 0.063 | 5.1 | 7.6 | 5.8 | +2.5 | +0.7 | Initial increase, peak at T3 (8.3%), then decline |
| **Stable Trajectories** |  |  |  |  |  |  |  |  |  |
| Problematic cannabis use | Intercept | 0.000 | -- | 10.3 | 10.5 | 9.9 | +0.2 | -0.4 | Stable |
| Problematic stimulant use | Intercept | 0.000 | -- | 3.4 | 3.4 | 4.2 | 0.0 | +0.8 | Stable |
| Problematic pornography use | Intercept | 0.000 | -- | 3.4 | 3.7 | 4.3 | +0.3 | +1.0 | Stable |
| **T2-T5 Trajectories** |  |  |  |  |  |  |  |  |  |
| Anxiety | Quadratic | 0.993 | 0.082 | — | 16.3 | 11.0 | — | -5.3 | Continuous curved decline |
| Depression | Quadratic | 1.000 | 0.007 | — | 9.1 | 6.1 | — | -3.0 | Continuous curved decline |

Note: T1 = April 2022 (pre-war); T2 = 2023 (immediately post-October 7, December 2023); T5 = February 2025. Piecewise models distinguish between Phase 1 (T1→T2, pre-war to immediate post-trauma) and Phase 2 (T2→T5, prolonged conflict period). Percentage in parentheses represents relative change from baseline.
<sup>a</sup> for model significance (F-test comparing to intercept model) across the whole time period

**Figure 1.**
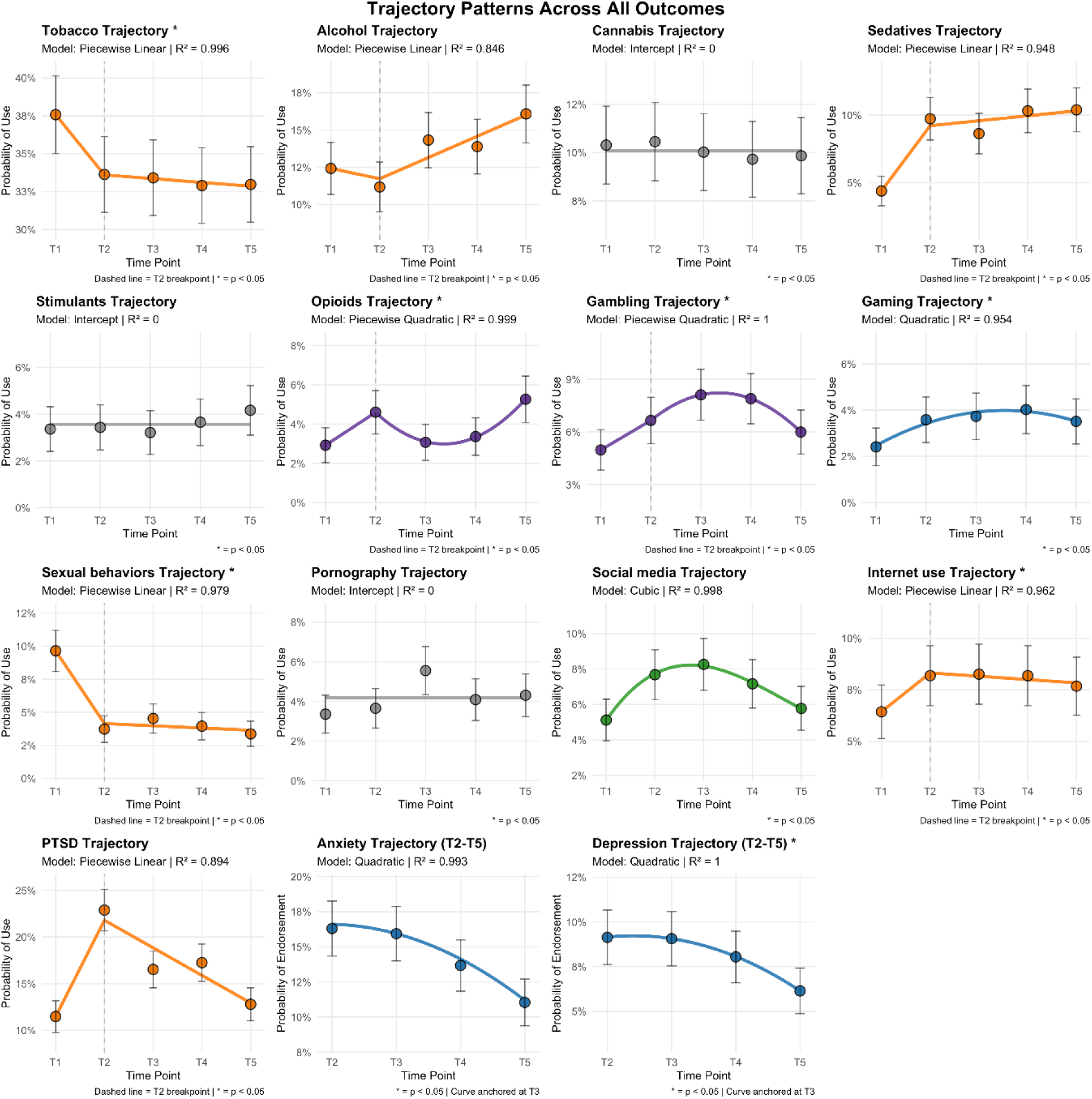
Trajectories of outcomes from pre-war (April 2022) to February 2025. Each panel displays observed prevalence (points with 95% confidence intervals) and best-fitting trajectory models (lines). Orange lines indicate piecewise linear models, purple lines represent piecewise quadratic models, blue lines show continuous quadratic trajectories, green lines show continuous cubic, and gray lines denote stable (intercept-only) patterns. T1=April 2022; T2=December 2023; T3=March 2024; T4=June 2024; and T5=February 2025. Vertical dashed lines at T2 represent the transition from pre-war (T1) to post-trauma (T2) and subsequent prolonged conflict period (T2-T5). Asterisks indicate statistically significant trajectories (p < 0.05) based on F-tests comparing to intercept.

#### Two-phase trajectories

Problematic tobacco use showed the most robust piecewise linear trajectory, with an initial -3.9% decrease from pre-war to immediate post-trauma (difference in prevalence between T2 and T1), followed by gradual stabilization during the prolonged conflict period (T2-T5), for an overall significant decrease (p=0.004). Conversely, problematic sedative use showed a sharp +5.3% rise immediately after October 7, with continued linear increase throughout the war period, although the overall change was not significant (p=0.052). Problematic use of alcohol showed an initial decrease from T1-T2, followed by linear increase; the overall increase (+3.7%) from T1-T5 was not significant (p=0.154). Problematic opioid use exhibited T1-T2 increase (+1.7%) followed by a quadratic (U-shaped) trajectory showing accelerating growth T2-T5, with an overall significant increase (p=0.03), suggesting compounding effects of prolonged conflict exposure. Problematic gambling showed T1-T2 increase (+1.7%) followed by a quadratic trajectory that was inverted U-shape, suggesting possible decreases towards T5, but with an overall significant increase (p<0.001). Problematic sexual behaviors demonstrated a sharp -5.9% drop T1-T2 and continued linear decline throughout T2-T5, with an overall significant decrease (p=0.021). Problematic internet use showed an initial +1.8% increase immediately after the trauma with stabilization from T2-T5, for an overall significant increase (p=0.038). PTSD showed an abrupt +11.4% surge immediately following the attacks (T1-T2), then gradually declined linearly T2-T5, for no overall significant increase (p=0.106).

#### Continuous and stable trajectories

Problematic gaming showed a steady quadratic increase, with prevalence gradually rising from 2.4% to 3.5% (p=0.046), possibly indicating progressive adaptation or cumulative stress. Problematic social media use showed a cubic trend, increasing from 5.1% to 8.3% at T3, then decreasing to 5.8%, with no overall significant change (p=0.063), suggesting possible habituation or changing coping mechanisms over time. From T2-T5, quadratic (curved) declines were observed for anxiety (−5.3%, but not statistically significant (p=0.082)) and depression (−3% decrease, p=0.007). The curved trajectory suggests rapid initial reduction followed by more gradual improvement, potentially reflecting psychological adaptation mechanisms or increased access to mental health services during the extended conflict period.

No significant change over time (intercept models) was observed for problematic cannabis use, problematic stimulant use, and problematic pornography use. These behaviors maintained stable prevalence rates despite the dramatic environmental changes, suggesting potential resilience.

### Moderation analysis

#### Gender

Gender significantly moderated trajectories for most outcomes (11/15) (Table 2). Women generally showed lower baseline prevalence for problematic use, and higher prevalence of internalizing disorders, but different patterns of change compared to men (Table 2; Figure 2). In some cases, different magnitude of change was observed for men and women. For problematic tobacco use, men showed a steeper immediate decline post-October 7 (from 46.9% to 40.2%) compared to women’s more modest decrease (28.1% to 26.9%). Similarly, problematic sexual behaviors demonstrated greater immediate decrease (−7.2%) in men than in women (−4.5%), and from T2-T5, men stabilized while women continued declining. Problematic alcohol use showed initial decreases which were greater in men than women, followed by parallel increases from T2-T5, maintaining men at higher prevalence. For anxiety, women (T2: 21.5%) started out higher than men (T2: 11.6%), and both genders showed linear declines across T2-T5, with steeper reduction in women and convergence by T5 (women: 11.5%, men: 10.7%). Similarly, women started with higher depression (T2: 10.3% vs. men: 8.3%) and both exhibited curved declines that were greater in women, with convergence by T5 (women: 5.9%, men: 6.5%).

**Table 2.** Gender moderation of trajectories.

| **Outcome** | **Best Model** | **Mod. p** | **Pseudo R<sup>2</sup>** | **Men T1→T5** | **Women T1→T5** | **Gap T1** | **Gap T5** | **Moderation Effect** |
| --- | --- | --- | --- | --- | --- | --- | --- | --- |
| Problematic tobacco use | Piecewise quadratic | <.001*** | 1.00 | 46.9%→39.2% (-7.7%) | 28.1%→26.5% (-1.6%) | 18.8% | 12.7% | Men declined more steeply than women; gap narrowed |
| Problematic gambling | Piecewise quadratic | <.001*** | 1.00 | 7.5%→9.6% (+2.1%) | 2.4%→2.4% (0.0%) | 5.1% | 7.2% | Men showed inverted-U pattern while women remained stable; gap widened |
| Problematic alcohol use | Piecewise linear | <.001*** | 0.99 | 17.9%→20.8% (+2.9%) | 6.8%→11.0% (+4.2%) | 11.1% | 9.8% | Phase 1 showed greater declines in men, with women showing greater overall increase. |
| Problematic pornography | Quadratic | .001** | 0.97 | 6.1%→7.5% (+1.4%) | 0.6%→1.1% (+0.5%) | 5.5% | 6.4% | Men showed curved pattern peaking at T3; women remained stable |
| Problematic sexual behaviors | Piecewise quadratic | .005** | 1.00 | 13.0%→5.8% (-7.2%) | 6.4%→0.9% (-5.5%) | 6.6% | 4.9% | Men showed larger Phase 1 decline |
| Problematic cannabis use | Piecewise quadratic | .011* | 1.00 | 12.3%→12.2% (-0.1%) | 8.2%→7.7% (-0.5%) | 4.1% | 4.5% | Differential intermediate fluctuations (T2-T4); women decreased slightly |
| Problematic stimulants use | Cubic | .013* | 1.00 | 4.6%→4.6% (0.0%) | 2.1%→3.6% (+1.5%) | 2.5% | 1.0% | Women increased while men remained stable; gap narrowed |
| Problematic gaming | Piecewise quadratic | .046* | 0.99 | 2.6%→3.0% (+0.4%) | 2.3%→3.9% (+1.6%) | 0.3% | -0.9% | Women increased more steeply; crossover (women surpassed men) |
| Problematic social media use | Quadratic | .033* | 0.93 | 3.6%→4.8% (+1.2%) | 6.8%→6.8% (0.0%) | -3.2% | -2.0% | Men increased while women remained stable; gap narrowed |
| Anxiety | Linear | .035* | 0.88 | 11.6%→10.7% (-0.9%) | 21.5%→11.5% (-10.0%) | -9.9% | -0.8% | Women showed steeper decline; convergence |
| Depression | Quadratic | .016* | 1.00 | 8.3%→6.5% (-1.8%) | 10.3%→5.9% (-4.4%) | -2.0% | +0.6% | Women showed steeper decline; crossover (men surpassed women) |

Note: In piecewise models, Phase 1 refers to T1 to T2; Phase 2 refers to T2 to T5. Changes shown as absolute percentage point change in parentheses. Negative gaps indicate women > men. For Anxiety and depression, change was T2→T5. Mod. = moderation \*p < 0.05, \*\*p < 0.01, \*\*\*p < 0.001

**Figure 2.**
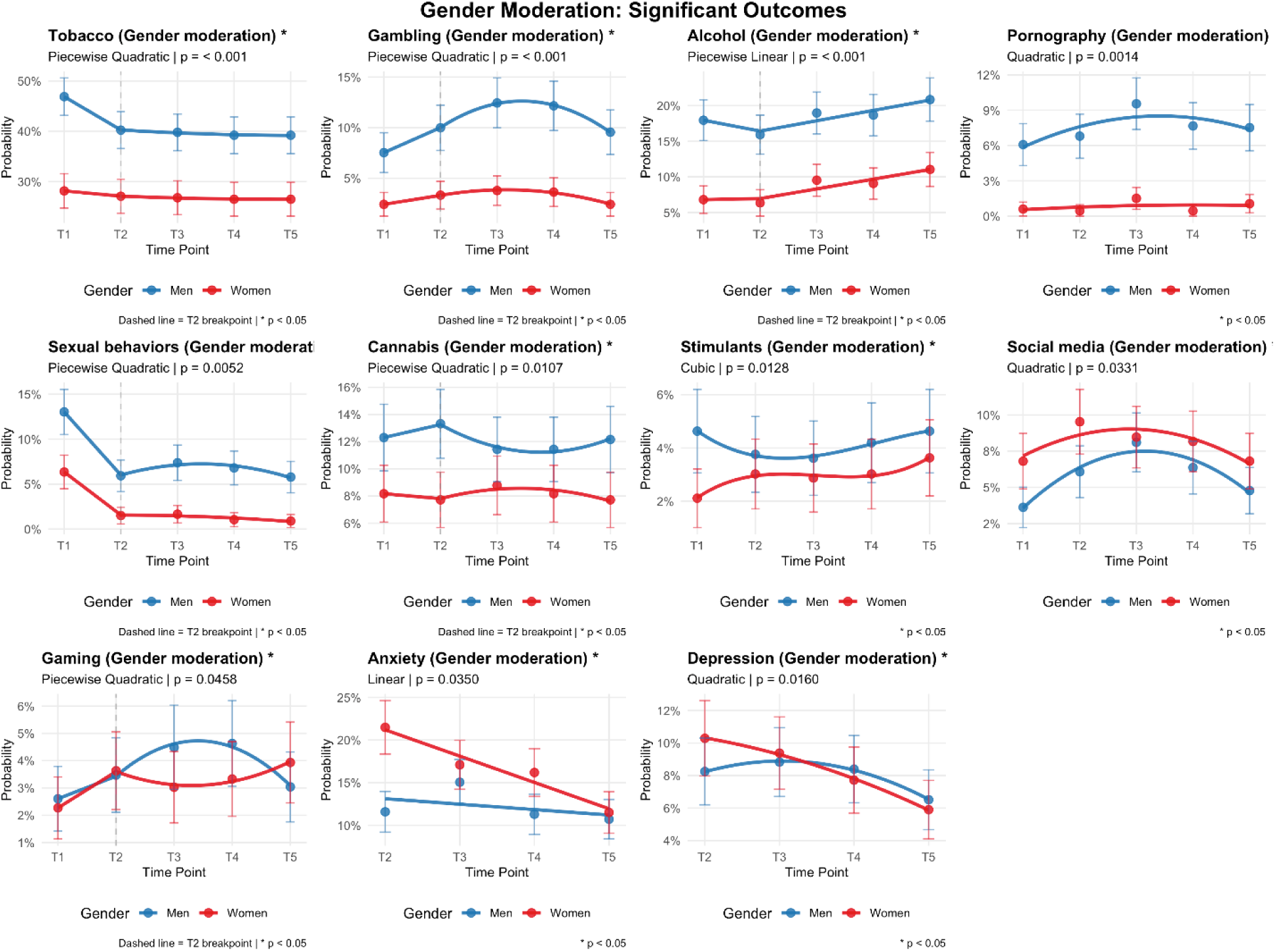
Gender moderation of longitudinal trajectories for significant outcomes. Displayed are predicted prevalence trajectories for outcomes exhibiting significant gender moderation effects across the study period. Trajectories are shown separately for men (blue) and women (red), with points representing observed prevalence and error bars denoting 95% confidence intervals. Solid lines represent model-based fitted trajectories based on the best-fitting functional form for each outcome (linear, quadratic, cubic, or piecewise models), as identified in Table 2. T1=April 2022; T2=December 2023; T3=March 2024; T4=June 2024; and T5=February 2025. Dashed vertical lines indicate the T2 breakpoint for piecewise models.

In other cases, change was observed in only one gender group. For problematic gambling, men showed a pronounced inverted-U pattern, peaking at T3 (13.3%) before declining, for a total increase of +2.1%, while women exhibited minimal change across all time points (remained at 2.4%). Problematic pornography use showed a curved pattern in men, peaking at T3 (9.6%) before declining, for an overall increase of +1.4%, while women maintained a stable low prevalence throughout (0.6% to 1.1%). Problematic stimulant use showed complex gender differences; overall, men remained stable while women showed significant increase of +1.5%. For problematic social media use, both genders exhibited a characteristic cubic pattern of initial increase followed by decline, for a net increase in men (3.6% to 4.8%) but no change in women (remained at 6.8%). Men and women showed different complex trajectories for problematic cannabis use. Men remained essentially stable (12.3% to 12.2%), while women showed a slight decline (8.2% to 7.7%). Complex patterns were also observed for problematic gaming, with men remaining stable overall (2.6% to 3.0%), and women increasing and becoming higher than men (2.3% to 3.9%).

Trajectories of change over time were not significantly moderated by gender for problematic use of: internet, opioids, sedatives; or PTSD.

#### Age

Age significantly moderated trajectories for all outcomes except problematic opioid use, demonstrating pervasive age-related differences in addiction vulnerability and response to conflict stress (Table 3; Figure 3). Notably, all significant age effects were negative (except problematic sedative use), indicating younger individuals (−1 SD age, approximately 29 years) generally showed higher prevalence and steeper increases during conflict compared to older adults (+1 SD age, approximately 56 years).

**Table 3.** Age moderation of trajectories.

| Outcome | Best Model | Mod. p | Pseudo R<sup>2</sup> | Age $\beta$ | Younger T1→T5 | Older T1→T5 | Moderation Effect |
| --- | --- | --- | --- | --- | --- | --- | --- |
| **Linear Trajectories** |  |  |  |  |  |  |  |
| Problematic gaming | Linear | <.001*** | 0.081 | -0.075 | 4.5%→7.2%<br>(+2.7%) | 0.7%→1.0%<br>(+0.3%) | Younger increased while older remained stable |
| Problematic pornography | Linear | <.001*** | 0.065 | -0.061 | 7.7%→9.5%<br>(+1.8%) | 1.1%→1.1% (0.0%) | Younger increased while older remained stable |
| Problematic cannabis use | Linear | <.001*** | 0.023 | -0.026 | 14.7%→16.5%<br>(+1.8%) | 4.7%→3.2%<br>(-1.4%) | Divergent directions: Younger increased, older decreased |
| Problematic alcohol use | Linear | <.001*** | 0.020 | -0.028 | 15.5%→19.8%<br>(+4.3%) | 9.2%→12.2%<br>(+3.0%) | Younger showed steeper increase |
| Problematic internet use | Linear | <.001*** | 0.089 | -0.073 | 17.2%→16.1%<br>(-1.1%) | 0.7%→1.1%<br>(+0.4%) | Divergent directions: Younger decreased slightly, older increased slightly |
| **Piecewise Linear Trajectories** |  |  |  |  |  |  |  |
| Problematic sexual behaviors | Piecewise linear | <.001*** | 0.073 | -0.065 | 10.8%→5.2%<br>(-5.6%) | 8.2%→1.0%<br>(-7.2%) | Older showed steeper phase 1 decline |
| Problematic tobacco use | Piecewise linear | <.001*** | 0.003 | -0.010 | 36.3%→33.7%<br>(-2.6%) | 35.1%→25.1%<br>(-10.0%) | Older showed steeper phase 1 decline |
| Problematic sedatives use | Piecewise linear | <.001*** | 0.016 | +0.011 | 3.7%→12.5%<br>(+8.8%) | 6.5%→14.3%<br>(+7.8%) | Younger showed steeper Phase 1 increase |
| PTSD | Piecewise linear | <.001*** | 0.035 | -0.029 | 20.9%→18.7%<br>(-2.2%) | 6.8%→8.6%<br>(+1.8%) | Younger steeper increase in phase 1 and steeper decrease in phase 2 |
| **Quadratic Trajectories** |  |  |  |  |  |  |  |
| Problematic gambling | Quadratic | <.001*** | 0.032 | -0.025 | 6.8%→9.0%<br>(+2.2%) | 3.3%→3.5%<br>(+0.2%) | Younger showed inverted-U pattern with net increase; older remained flat |
| Problematic stimulants use | Quadratic | <.001*** | 0.038 | -0.031 | 4.8%→5.5%<br>(+0.7%) | 2.1%→2.8%<br>(+0.7%) | Similar absolute change but different curve shapes; younger flatter, older more curved |
| Problematic social media use | Quadratic | <.001*** | 0.070 | -0.100 | 11.7%→14.3%<br>(+2.6%) | 0.7%→0.7% (0.0%) | Younger showed dynamic inverted-U pattern (peaked at T3); older remained stable |
| Anxiety | Linear | <.001*** | — | -0.034 | 24.0%→16.1%<br>(-7.9%) | 12.4%→7.6%<br>(-4.8%) | Younger showed steeper absolute decline |
| Depression | Linear | <.001*** | — | -0.037 | 15.1%→8.7%<br>(-6.4%) | 5.0%→3.5%<br>(-1.5%) | Younger showed steeper absolute decline |

Note: Age centered at mean (42.5 years); -1SD ≈ 29 years, +1SD ≈ 56 years. In piecewise models, Phase 1 refers to T1 to T2; Phase 2 refers to T2 to T5. Changes shown as absolute percentage point change in parentheses.. $\beta$ = log-odds change per SD increase in age. For Anxiety and depression, change was T2→T5. Mod. = moderation \*p < 0.05, \*\*\*p < 0.001

**Figure 3.**
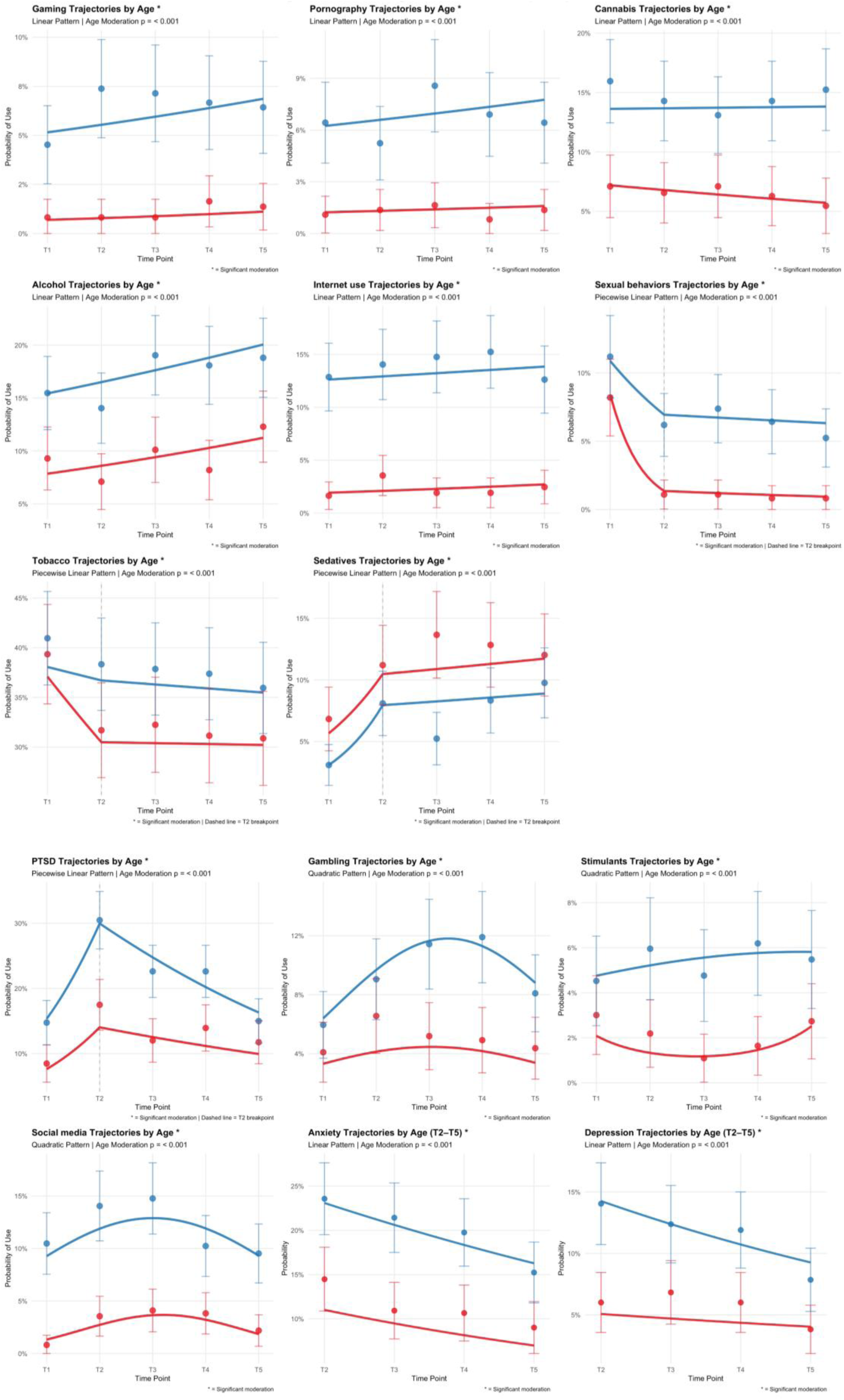
Age moderation of longitudinal trajectories for significant outcomes. Predicted probabilities are shown separately for younger adults (−1 SD age; blue) and older adults (+1 SD age; red). Solid lines represent model-estimated trajectories, with points indicating observed prevalence at each wave and error bars denoting 95% confidence intervals. T1=April 2022; T2=December 2023; T3=March 2024; T4=June 2024; and T5=February 2025. Vertical dashed lines indicate the T2 breakpoint for piecewise linear models.

In some cases, change was only observed for younger individuals, while older adults remained stable. Problematic gaming showed the strongest age effect (β = -0.075), with younger adults significantly increasing from 4.5% to 7.2%. For problematic gambling, younger adults showed an inverted U shape, peaking at T3 (11.8%) before declining to 9.0%. Similarly, for problematic pornography use, younger adults showed an increase (+1.8%, from 7.7% to 9.5%). Last, for problematic social media use, younger adults showed a dynamic pattern with initial increase (11.7% to peak of 19.4% at T3) followed by decline (to 14.3% at T5).

In other cases, differential change was observed. For problematic alcohol use, younger adults showed a steeper increase (15.5% to 19.8%) than older adults (9.2% to 12.2%). For problematic tobacco use, both age groups decreased from T1 to T2, but older adults exhibited a steeper decline (35.1% to 26.5%, -8.6%) compared to younger adults’ more modest decrease (36.3% to 34.1%, -2.2%). Problematic cannabis use showed opposing directions of change, with slight increase in younger adults (14.7% to 16.5%, +1.8%) and slight decrease in older adults (4.7% to 3.2%, -1.4%). For problematic sedative use, from T1 to T2, younger adults demonstrated a steeper rate of increase (+5.9%) compared to older adults (+4.7%). For problematic stimulant use, younger adults showed a flatter trajectory compared to older adults’ inverted U-shape pattern.

For problematic sexual behaviors, younger adults showed a sharp Phase 1 decline (10.8% to 6.2%) followed by a continued decrease to 5.2%, while older adults showed an even steeper initial decline (8.2% to 1.3%) with maintenance at minimal levels (1.0%). For problematic internet use, both groups showed modest fluctuation with different patterns (younger decreased, older increased), with the large age gap persisting across all time points. For PTSD, during Phase 1 (T1→T2), younger adults showed a dramatic spike (+14.3%) compared to older adults’ more modest increase (+6.1%); during Phase 2 (T2→T5), both groups declined, but younger adults showed a steeper recovery (−16.5%) compared to older adults (−4.3%). Last, younger adults showed steeper declines for anxiety and depression.

## Discussion

In a longitudinal study of an adult Jewish general population sample from Israel, with data from before the October 7^th^ terror attacks (April 2022) to immediately after (December 2023) and over the course of the subsequent war (March 2024, June 2024, and February 2025), we observed different patterns of change in the prevalence of problematic substance use and behaviors and common mental health issues. For some outcomes, different trajectories were observed before and after December 2023, suggesting that the events of October 7^th^ and the early war may have been a key transition point. Other outcomes showed smooth changes over time, while still others did not change significantly. Additionally, many outcomes showed different trajectories by gender and age. This information is important for exploring possible effects of trauma and war, identifying outcomes requiring attention, and suggests that outcome-specific and group-specific strategies may be warranted.

Overall, the high model fits achieved suggest that population-level trajectories followed predictable patterns despite individual variability, supporting the utility of such modeling for understanding collective psychological responses to mass trauma events. Specifically, outcomes that fit best to a piece-wise model suggest proximal responses. For example, the sharp increase in high levels of trauma-related (PTSD) symptoms followed by gradual decline over time was also observed in previous studies^16,22,23^, as would be expected following mass trauma. Similarly, the decreases in anxiety and depression over time are expected, as people adapt and cope with their feelings. However, the apparently elevated T2 starting points suggest substantial acute distress immediately following the attacks, with subsequent improvements potentially reflecting regression to pre-conflict baselines or establishment of a “new normal” that remains elevated above peacetime levels. Furthermore, psychological distress (PTSD, anxiety, depression) was higher in the younger age group, similar to previous studies worldwide^64,65^ and after October 7^th^ in Israel^12,13,27^, and both increased (for PTSD) and then decreased at sharper rates, indicating more stress reactivity and emotional lability. Generally, women are considered to be at higher risk for depression and anxiety^66–68^, and that was observed in Israel closer to the beginning of the war^16,17,25^. In our study, prevalence appeared to converge for women and men over time, similar to what was observed for depression previously^66^. Additional studies are needed to better understand the complex mechanisms underlying emotional and psychological responses to trauma and increased vulnerability to distress, and why men may be responding differently more recently.

While some previous studies showed increases in substance use post-October 7^th^ ^13,18,20^, this is the first study to explore changes in problematic substance use. A proximal increase in problematic use of sedatives was observed, perhaps indicating self-medication to cope with the traumatic events, with more gradual increase over the course of the war, perhaps due to ongoing stress. This can be especially problematic, because using sedatives (such as benzodiazepines) after trauma was associated with increased risk for PTSD^69^. Problematic use also increased for other prescription medications (opioids and stimulants (in women)), suggesting that people may turn to what is familiar to them, but in a non-medical way. Clinicians should discuss with patients the possible harms of misusing prescription medications and suggest more adaptive methods of coping with trauma and stress. While problematic alcohol use initially decreased, which may happen during acute stress as people want to be more fully aware, or may be due to less social drinking due to the war, prevalence then began to increase, as people returned to baseline or used alcohol more to cope. Surprisingly, problematic tobacco use decreased, especially in men and older adults, but remained the most prevalent outcome. Additional studies should identify factors underlying the decrease, to precipitate further declines.

In terms of problematic behaviors, the only previous study we are aware of used data from the same general sample as this (through June 2024), and showed that problematic gambling increased among men with difficulties in emotional regulation^29^. Here, curved increases were observed for problematic gambling (specifically among men and younger adults) and gaming (specifically among women and younger adults) suggesting group-specific dynamic reactions to stress, which should be studied further. Problematic internet use also increased proximally (after October 7^th^), perhaps as people increasingly used internet to follow events, and then leveled off to a new normal. Similarly, problematic use of social media increased in the beginning, especially in younger adults, but then began to decrease, perhaps due to people realizing that such use may increase their exposure to potentially harmful or traumatic content, such as graphic media or hate speech^70^.

The implications of these findings on potential effects of ongoing terror and war on mental health are discussed. First, ongoing monitoring is needed to have consistent epidemiological data on the prevalence of mental health issues and problematic substance use and behaviors, to be aware of and responsive to changes that may occur. Second, public health strategies could be developed to address two key issues. One, to educate that using substances or behaviors to cope with stress might appear to work in the short term, but may lead to longer term harm, and may prevent the inherent resilience mechanisms in the body from working properly. Two, to provide discussion about alternate, more beneficial ways to cope with ongoing stress and increase resilience. Third, targeted interventions should be developed for vulnerable groups (e.g., young adults), and further research is needed to more fully understand the risk factors for each outcome.

## Limitations

Study limitations are noted. First, this study only includes Jews, and those who were part of iPanel. Culturally-appropriate, large-scale studies in the Arab population of Israel, which may have been more affected by trauma and stress^11,15^, are needed. Second, only a subset of the samples agreed to participate at the different time points. We used multiple imputation to deal with missing data, but how generalizable the results are is unknown. In April 2022, there were differences between those who participated in at least one additional survey and were included in the longitudinal study (n=1,368) and those who were not (n=1,291). Those included were significantly older than those who were not (p<0.0001); after adjusting for the age affect, those included differed from those not included for education (p=0.007) and self-perceived economic level (p=0.026), but did not differ significantly in terms of mental health issues (problematic substance use and behaviors, PTSD symptoms, and difficulties in emotion regulation). Third, we looked at prevalence for dichotomous outcomes, which are dependent on a specific threshold, but important for public health. Fourth, change might have happened before October 7^th23^. Additional studies that explore if trajectories differed by October 7^th^ exposure may better address the source of the changes. Fifth, self-report, retroactive measures could lead to recall and other biases, although validated measures were used. Sixth, since the war-related trauma was ongoing and respondents may not necessarily be post-trauma, we are not formally assessing PTSD, but rather PTSD symptoms due to trauma and possibly ongoing stress. Follow-up studies after cessation of traumatic events will provide more precise estimates of formal PTSD.

## Conclusions

This study provides key public health information about trajectories over time in the prevalence of problematic substance use and behaviors and mental health issues in Israel from before October 7^th^ and over the course of the war, to suggest possible psychological, emotional, and behavioral reactions to trauma and ongoing stress. On both the population and individual levels, it is important to be aware of the potentially harmful effects of such life events, as well as the consequences of maladaptive coping styles on health and well-being.

## Data Availability

All data produced in the present study are available upon reasonable request to the authors

---

**Supplemental Table 1.** Number of respondents at each time point

| Collection dates | April 2022 | December 2023 | March 2024 | June 2024 | February 2025 |
| --- | --- | --- | --- | --- | --- |
| 2022 | **2659** | 1343 | 1053 | 1052 | 839 |
| 2023 |  | **4002** | 2768 | 2757 | 1998 |
| 2024 March |  |  | **2768** | 2270 | 1676 |
| 2024 June |  |  |  | **2757** | 1747 |
| 2025 |  |  |  |  | **2912** |

**Supplemental Table 2.**
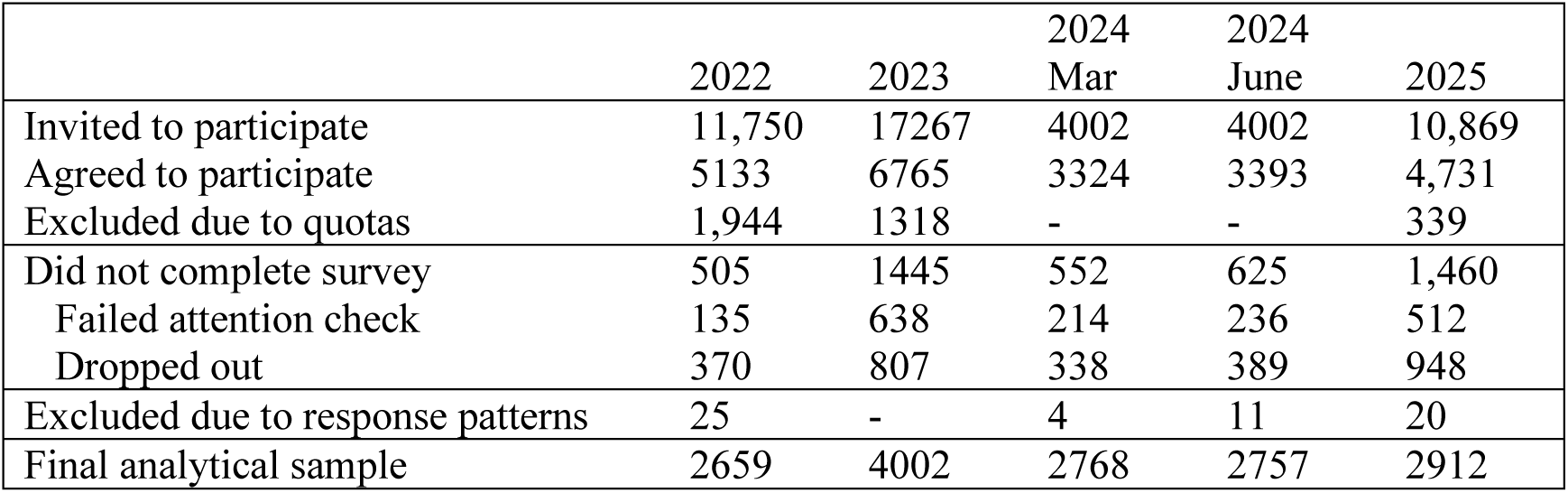
Information on survey response and completion rates

|  | 2022 | 2023 | 2024 Mar | 2024 June | 2025 |
| --- | --- | --- | --- | --- | --- |
| Invited to participate | 11,750 | 17267 | 4002 | 4002 | 10,869 |
| Agreed to participate | 5133 | 6765 | 3324 | 3393 | 4,731 |
| Excluded due to quotas | 1,944 | 1318 | - | - | 339 |
| Did not complete survey | 505 | 1445 | 552 | 625 | 1,460 |
| Failed attention check | 135 | 638 | 214 | 236 | 512 |
| Dropped out | 370 | 807 | 338 | 389 | 948 |
| Excluded due to response patterns | 25 | - | 4 | 11 | 20 |
| Final analytical sample | 2659 | 4002 | 2768 | 2757 | 2912 |

**Supplemental Table 3.** Sample descriptives (based on Time 1; N=1368)

|  | *n* | *Percent (%)* |
| --- | --- | --- |
| **Gender** |  |  |
| Men | 691 | 50.5 |
| Women | 661 | 48.3 |
| Unknown | 16 | 1.2 |
| **Age** |  |  |
| 18-25 | 164 | 12.0 |
| 26-34 | 304 | 22.2 |
| 35-49 | 440 | 32.2 |
| 50-70 | 460 | 33.6 |
| **Religiosity** |  |  |
| Secular | 681 | 49.8 |
| Traditional | 394 | 28.8 |
| National Religious | 162 | 11.8 |
| Ultra-Orthodox | 131 | 9.6 |
| **Residence Area** |  |  |
| Jerusalem Area | 160 | 11.7 |
| Tel Aviv / Center area | 656 | 48.0 |
| Haifa / North | 344 | 25.1 |
| South | 166 | 12.1 |
| Yehuda & Shomron | 42 | 3.1 |
| **Educational attainment** |  |  |
| High school, non-matriculation | 108 | 7.9 |
| High school, matriculation | 243 | 17.8 |
| Post-high school, technical | 293 | 21.4 |
| Post-high school, academic | 724 | 52.9 |
| **Ethnicity** |  |  |
| Middle Eastern/North African | 522 | 38.2 |
| European, not Former Soviet Union | 466 | 34.1 |
| Former Soviet Union | 123 | 9.0 |
| Mixed | 226 | 16.5 |
| Other | 31 | 2.3 |
| **Marital status** |  |  |
| Single | 398 | 29.1 |
| Married | 853 | 62.4 |
| Separated/Divorced/Widowed | 117 | 8.6 |
| **Socio-economic status – subjective** |  |  |
| Much below average | 123 | 9.0 |
| Below average | 125 | 9.1 |
| Slightly below average | 157 | 11.5 |
| Average | 547 | 40.0 |
| Slightly above average | 283 | 20.7 |
| Above average | 109 | 8.0 |
| Much above average | 24 | 1.8 |

**Supplemental Figure 1.**
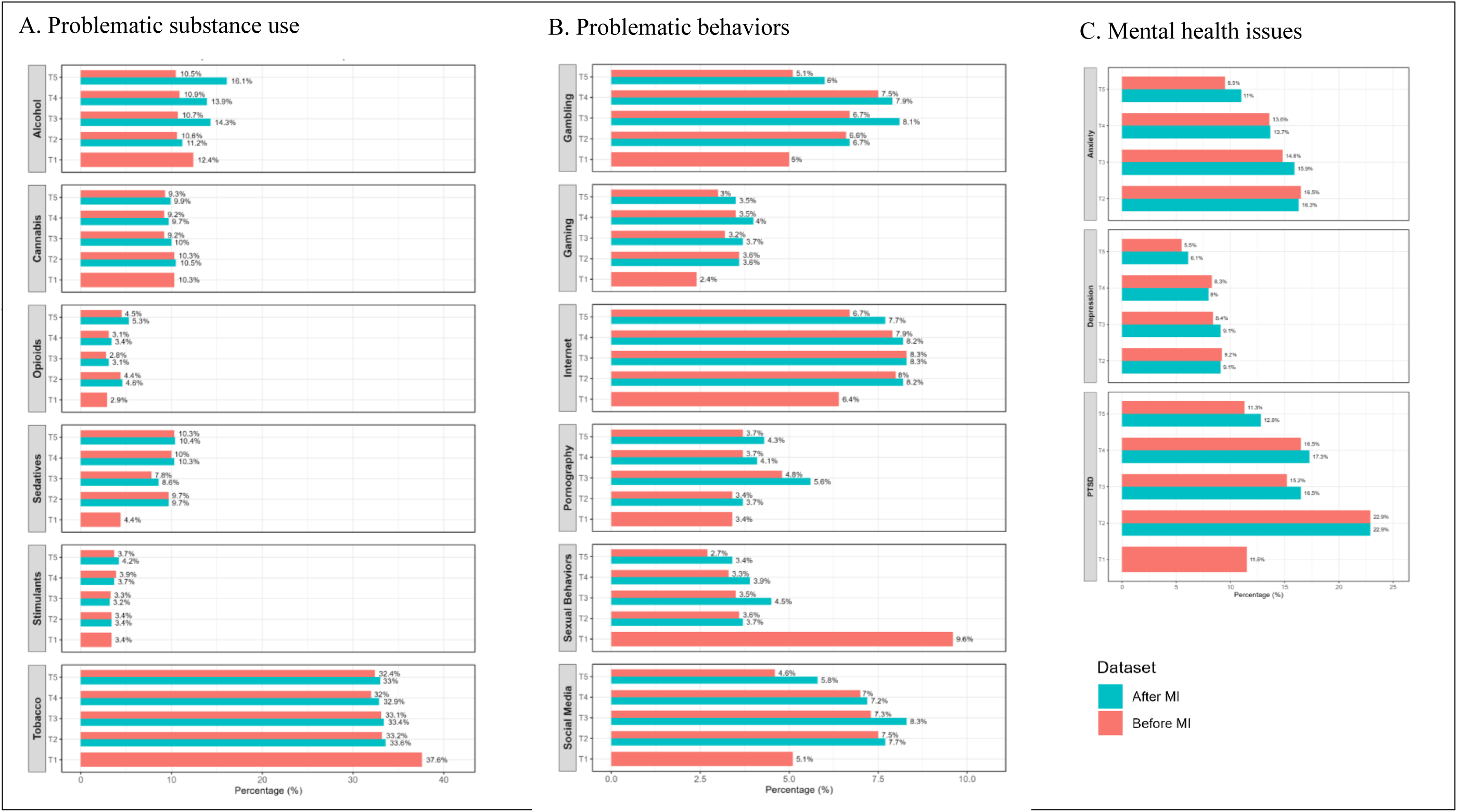
Prevalence of outcomes over all time points (N=1,368)

